# Predictors of post-stroke cognitive impairment at three-month following first episode of stroke among patients attended at tertiary hospitals in Dodoma, central Tanzania: a protocol of a prospective longitudinal observational study

**DOI:** 10.1101/2022.08.05.22278475

**Authors:** Baraka Alphonce, John Meda, Azan Nyundo

## Abstract

**Introduction:** Neurocognitive deficits after stroke are a common manifestation and pose a significant impact on the quality of life for patients and families; however, little attention is given to the burden and associated impact of cognitive impairment following stroke. The study aims to determine the prevalence and predictors of post-stroke cognitive impairment (PSCI) among adult stroke patients admitted to tertiary hospitals in Dodoma, Tanzania

**Methodology:** A prospective longitudinal study is conducted at tertiary hospitals in the Dodoma region, central Tanzania. Participants with the first stroke confirmed by CT/MRI brain aged ≥ 18 years who meet the inclusion criteria are enrolled and followed up. Baseline socio-demographic and clinical factors are identified during admission, while other clinical variables are determined during the three-month follow-up period. Descriptive statistics are used to summarize data; continuous data will be reported as Mean (SD) or Median (IQR), and categorical data will be summarized using proportions and frequencies. Univariate and multivariate logistic regression analysis will be used to determine predictors of PSCI

## Introduction

Stroke is the leading cause of death and disability; approximately sixty-seven million people worldwide suffer their first stroke yearly, of which close to six million die and five million live with associated disabilities (1,2). The incidence of stroke often presents diverse outcomes, including neurocognitive impairment, which significantly impacts the quality of life, increased health care cost, loss of income, and social isolation for the patients (3–5).

PSCI prevalence ranges from 35 to 92 percent (6–8); among the few studies conducted in Sub-Saharan Africa, a three months prevalence of 40% was reported in Nigeria and a year prevalence of 34% was observed in Ghanaian stroke survivors (9,10). Several factors may contribute to the discrepancy in prevalence across settings, differences in the screening and diagnostic tools used to assess PSCI in different studies such as MoCA, MSSE, or neuropsychological test could partly explain the phenomenon, furthermore, the timing of screening for cognitive impairment following a stroke, ethnicity, and cultural and educational backgrounds may also predict the differences(11)

While an episode of stroke is an independent predictor for cognitive impairment, specific patient may play a role in the development of PSCI. These include, advanced age, female gender, fewer years of formal education, alcohol consumption, and cigarette smoking have been linked to cognitive impairment after stroke. Cardiometabolic conditions such as, hypertension, diabetes, dyslipidemia, atrial fibrillation, stroke-specific characteristics, and neuropsychiatric symptoms at baseline have all been linked PSCI(12–17).

Therefore, the objective of the study is to assess the prevalence and predictors of early post-stroke cognitive impairment after the first episode of stroke among patients admitted at tertiary hospitals in Dodoma, Tanzania

## Materials and Methods

### Study aims

1. To determine the baseline prevalence of cognitive impairment at one month following the first stroke among adult patients admitted at tertiary hospitals in Dodoma, Tanzania.
2. To determine the cumulative incidence of cognitive impairment at three-month following the first stroke among adult patients admitted at tertiary hospitals in Dodoma, Tanzania.
3. To determine the predictors of post-stroke cognitive impairment at three-month after the first stroke among adults admitted at tertiary hospitals in Dodoma, Tanzania.

### Study design

A prospective longitudinal observational study

#### Study setting

This study is currently ongoing in the Internal Medicine Departments of the Dodoma Regional Referral Hospital (DRRH) and Benjamin Mkapa hospital (BMH), with a bed capacity of 480 and 400, respectively. The two hospitals cover the estimated 2.5 million population in the Dodoma region as per the 2012 census (18). The hospitals also serve the neighboring districts of Singida, Iringa, Manyara, and Morogoro regions. According to local statistics, stroke is one of the leading causes of admission to referral hospitals in Dodoma, accounting for 15 percent of all monthly admissions. In addition, Dodoma regional Referral Hospital and Benjamin Mkapa Hospital are the designated teaching hospital for the University of Dodoma for medical training in both undergraduate and residency programs. With its well-built infrastructure, the Benjamin Mkapa Hospital is equipped with neuroimaging services including CT scans and MRI serving Dodoma and the central part of Tanzania as a whole.

#### Sample size estimation

The sample size will be estimated utilizing the Krejcie and Morgan formula for prospective studies(19)

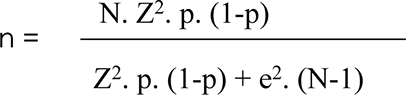

n - Calculated sample size

N - Population sample size from the previous study

Z - Confidence interval level

p - The true probability of an event

e - Margin error

N - 140 (20)

Z - Confidence interval at 99% (standard value of 2.56)

P - 39.9% (20)

e - Marginal error of 0.01

n = 180

#### Inclusion criteria

1. Patients aged 18 years and above who are admitted with the first episode of stroke
2. Able to provide informed consent or a proxy consent from a close relative in case the patient is incapable
3. Adults patient with first-ever stroke presenting within 14 days and confirmed by CT scan or MRI brain

#### Exclusion criteria

1. Patient with traumatic intracerebral hemorrhage
2. Patient with intracerebral hemorrhage due to tumor
3. Patient with severe sensory impairment (blindness and deafness)
4. Patient with Transient Ischaemic Attack (TIA)
5. Patient with subarachnoid hemorrhage
6. Patients with previous neurological disorders such as epilepsy

### Outcome measurement

#### Primary outcome

The outcome of interest is post-stroke cognitive impairment, which will be evaluated by a certified investigator one month after stroke onset as a baseline and again three months later (22). The cognitive impairment will be assessed using Montreal cognitive assessment (MoCA) tools, version 7.0, but participants with less than 12 years of education will receive an additional point. The cognitive impairment will be set at a cutoff score of 23 out of 30 (23). The cognitive decline will be defined as a reduction in MoCA score by two or more points between baseline assessment at one month and the second evaluation at three months, and cognitive improvement as an increase of two or more points (24)

##### Aim 1 Study Variables

the variables address the prevalence of PSCI at one month and three months using MoCA, which constitutes key cognitive domains including visual-spatial executive functioning, sentence repetition, and a phonemic fluency task, short-term memory, abstraction, attention and calculation, and orientation (time and space) (See table 1 for a listing and brief explanation of the variables for the Aim 1).

##### Aim 2 Study Variables

Assess the unit decline of cognitive functioning from the baseline to the three-month point after the stroke episode. A minimum of two or more points decline in standardized MoCA between the time intervals will be considered a significant decline (24). (See table 3 for a listing and a brief explanation of the variables for Aim 2

##### Aim 3 Study Variables

The variables address the predictors of PSCI at three months; these include Age (in years), sex, alcohol use, smoking history, history of diabetes mellitus, dyslipidemia, atrial fibrillation, post-stroke depression, apathy, stroke type and characteristic (hemorrhagic/ischaemic, cortical/sub-cortical), stroke (infarct/hematoma) volume, presence of Leukoaraiosis or brain atrophy (See table 4 for listing and a brief explanation of the variables for the Aim 3)

#### Secondary outcome

Death will be recorded as a secondary outcome; this data will be collected from hospital records and through contact with study participant’s next of kin; the length of time between hospitalization and death will be determined.

#### Participants’ characteristics

Participants are adults aged 18 years or older admitted to the Internal Medicine wards of either DRRH or BMH with a diagnosis of the first stroke as per the World Health Organization, defined as “rapid development of clinical signs of focal or global disturbance of cerebral function lasting more than 24 hours or leading to death, with no apparent cause other than vascular origin” and confirmed by CT scan or MRI, with the duration of symptoms lasting not more than 14 days (21).

### Data collection process

#### Evaluation of the participants

As it stands, 100 patients who meet the inclusion criteria have been enrolled since June of 2021. Participants who provided informed consent are interviewed using a structured questionnaire that collects demographic data such as age, gender, and level of education. A minimum of two contacts, one of the patient’s and that of the next of kin, are recorded. Information regarding potential predictors of PSCI are also collected, these include, alcohol consumption history (defined as alcohol consumption within the past 12 months), with an emphasis on duration and frequency (25,26). Duration and number of cigarettes packs per day, the history of current smoking, defined as smoking within the past year, is also obtained (27). A history of Hypertension (defined as a history of Hypertension or the use of antihypertensive medications) and diabetes (defined as a history of diabetes or the use of diabetic medications) are documented (28)

### Clinical variable measurement

#### Post stoke cognitive impairment

At one and three months after a stroke, Montreal cognitive assessment (MoCA) measures cognitive impairment; participants with a MoCA score of < 23/30 are considered to have screened positive for cognitive impairment (29). Compared to an initial cutoff score of 26, a MoCA score of 23 is considered more sensitive and lowers the false positive rate and exhibits overall improved diagnostic accuracy(30). The use of MoCA has been verified in Tanzania among the older population, a score of 22 indicates a cognitive impairment with a sensitivity of 90% and specificity of 80% (28).

Given that the MoCA scale is less sensitive to milder forms of cognitive impairment and susceptible to low educational levels, an additional point is usually added for those with less than 12 years of formal education (32). The use of MoCA instead of the ideal comprehensive neuropsychological battery may reduce the diagnostic accuracy, MoCA remains among few reliable scales in screening of cognitive functioning in a pragmatic clinical settings, indeed, MoCA assess cognitive domains that reflects similar constructs to the measurements of comprehensive neuropsychological battery (33)

#### Post-stroke depression

The Patient Health Questionnaire (PHQ) – 9 with a total score of 27 is used to screen stroke survivors for post-stroke depression; a score of 1 – 4 is considered minimal depression, 5 – 9 as mild depression, 10 – 14 as moderate depression, 15 – 19 as moderately severe depression, and 20 – 27 as severe depression. At a PHQ score of 15, the likelihood of having a major depressive disorder is marked. (Kroenke, Spitzer, & Williams, 2001). In studies, the diagnostic validity of the 9-item PHQ-9 has been proven for Major Depressive Disorder; a PHQ-9 score of 10 indicated an 88 percent sensitivity and an 88 percent specificity (34). Individuals with a low PHQ-9 score of 4 had a 1 in 25 risk of becoming depressed (35). A study done in Tanzania proved that the PHQ-9 is the right tool for screening depressive symptoms equivalent to a major depressive episode in primary health care settings. The results favor a cutoff score of ≥ 9, with a sensitivity of 78% and a specificity of 87% for depressive symptoms equivalent to a major depressive episode (36)

#### Post-stroke apathy

Apathy is evaluated at one month using the apathy evaluation scale; those who met the criteria scored more than 38. This cutoff score (EAS > 38) has sensitivity and specificity of around 80 % and 100%, respectively (37), to identify suggestive apathy symptoms (38)

#### Blood pressure measurement

A blood pressure (BP) reading are measured using an automated digital machine of AD Medical Inc. brand, keeping with the 2018 AHA/ACC Hypertension guideline for standard measurement of BP (39). The affected arm is avoided to minimize false readings from the impact of muscle tone changes; Hypertension will be defined as BP ≥140/90 mmHg after three different reading measured at least 5 minutes apart or a patient with a known history of hypertension or a patient on antihypertensive medications (40).

#### Radial pulse measurement

Two operators assess the radial pulse measurement; “Operator 1” listens to the heart sounds with a stethoscope, while “operator 2” palpates the radial pulse while the patient is seated. When both operators are ready, a timer will be utilized to begin counting simultaneously. The technique is repeated, and the heart rate is recorded after 1 minute. To eliminate bias, the two operators switch roles on different patients. The difference between the rates counted by the two operators at the end of one minute are used to determine the apex-pulse deficit (41). A deficit of ten or more is considered indicative of atrial fibrillation (42). In addition, each operator assigns a regular or irregular rhythm to the beat

#### Stroke severity

On the day of enrollment, stroke clinical presentations is recorded, and the NIHSS will be used to categorize stroke severity as minor, mild, moderate to severe (World Health Organization, 2010)

### Laboratory investigations

#### Collection of blood sample

The laboratory works for the study are based on standard operating procedures from credited Dodoma Regional Referral Hospital. Each participant will be asked to consent to venipuncture and finger prick then a laboratory technician follows the aseptic procedure before taking blood samples. Before sample collection, tubes [EDTA (K2/K3) sodium fluoride plain, no additives] are labeled with the hospital registration number corresponding to the patient then a tourniquet is placed proximally on the upper arm approximately 60cm above the elbow joint. The intended site of venipuncture is cleaned with 70% methylated spirit in an enlarging circulatory pattern, and the area is left to dry for 20 seconds. Then 10mls of venous blood are taken from each arm using a 5cc syringe for lipid profile (HDL, LDL, Total Cholesterol, and Triglycerides). Then the tourniquet is removed, and the site of venipuncture pressed with a cotton swab to arrest bleeding. After disposing of the syringes and using gloves, the blood samples are transported into a cool box to a credited DRRH laboratory within three hours of sample collection. However, hemolyzed blood is not be processed rather venipuncture will be repeated. The serum sample is separated from whole blood by centrifugation at 300 rpm for 5 minutes, and two aliquots are prepared, one for lipid profile and another for serum electrolytes. A sample is stored at 2^0^C – 8^0^C if the analysis is expected after 2 hours from sample collection; however, blood samples are stored at room temperature if the analysis is expected to be done within 2 hours from sample collection. Analysis of the sample is done using clinical chemistry automatic analyser Erba machine XL – 180 serial number 160239 made in Germany.

For dyslipidemia: is defined as a high total cholesterol level above 200 mg/dL, or a low-density lipoprotein cholesterol level above 130 mg/dL, or a triglyceride level above 150 mg/dL, or a high-density lipoprotein cholesterol level below 40 mg/dL for women and below 50 mg/dL for men(10).

Blood Sugar measurement: a blood sample for measuring blood sugar is obtained from a finger prick with the guidance of sample collection manual SM-1-03.3 of DRRH laboratory. After the candidate’s consent, the palm is positioned side up, and then the fingertip of the index, ring or middle finger is chosen; then, pressure is applied on the finger to increase blood flow to the fingertip. The fingertip is cleaned with methylated spirit 70% from the middle to the periphery after 15 minutes the finger will be dry. Using a new sterile lancet placed firmly against the periphery of the fingertip to puncture the skin to increase blood flow, the finger is held below the level of the elbow. Then the blood sample collected directly from the device (ACCU-CHECK Active Roche glucometer machine). After collecting the sample, the client is given a ball of cotton wool to press against the finger for 10 minutes to arrest bleeding, and the lancet will be disposed to a sharp disposal box. Hyperglycemia is defined according to American Diabetes Association (43); for non-diabetic patients, hyperglycemia is defined as random blood sugar >11.1 mmol/L or fasting blood sugar > 7.0 mmol/L, and diagnosis of diabetes will be made with a fasting blood sugar ≥ 7.0 mmol/L, or random blood glucose ≥ 11.1 mmo/L plus symptoms of hyperglycemia or glycated hemoglobin≥ 6.5 % To ensure validity and reliability of the results, the laboratory technician in respective hospitals, the Benjamin Mkapa Hospital and Dodoma regional referral hospital, tends to run controls in every machine every day before processing samples, and maintenance of machines is done weekly, whereas critical services are done in every six months

#### Electrocardiogram

The principal investigator, familiar with the manufacturer’s instructions, uses a 12-lead ECG (model C12G, manufactured by ARI Technology Medical Co. Ltd 2019) on each participant under the supervision of a consultant cardiologist. Before a 12-lead ECG is recorded, the patient is informed about the procedure, privacy is respected in a comfortable environment enhance relaxation and reduce interference with the ECG trace’s clarity. To attach the electrodes/leads to the patient, the appropriate equipment, such as the electrocardiograph, ECG paper, and ECG tabs, is provided. Ensuring that the ECG cables are not twisted is critical, leading to ECG tracing interference. The patient’s ID number is entered into the machine, consent obtained, and a patient is requested to lie down at an angle of 45^0^ with the head adequately supported and the backrest on the bed, with the inner aspect of the patient’s wrist close to but not touching his or her waist.

Provided that wet gel electrodes are utilized, there is no need to shave the skin; then, limb electrodes will be applied in the following order: red on the right inner wrist, yellow on the left inner wrist, black on the right inner leg just above the ankle, green on the left inner leg just above the ankle. The following is how the chest leads are organized: V1 is just to the right of the sternum, V2 is just to the left of the sternum, V3 is halfway between V4 and V2, V4 is in the fifth intercostal space, mid clavicular line, V5 is in the anterior axillary line, and V6 is in the mid axillary line, the same horizontal line as V4 and V5. ECG cables should not lie to each other, and tension should be avoided to decrease artifacts and increase the accuracy and quality of ECG tracing. The calibration signal on the ECG machine should be kept at a paper speed of 25 millimeter/second and

ECG size 1 millivolt/10-millimeter deflection. The patient is requested to lie still and breathe regularly during the operation; the ECG trace should be clear before recording. The patient’s name, hospital number, date of birth, and the day and time the ECG was obtained are be written on the 12-lead ECG trace (44). According to the American College of Cardiology guideline for the management of patients with atrial fibrillation, Atrial Fibrillation will be diagnosed by the absence of P waves and irregular-irregular RR interval (45)

With a 24-hour ECG Holter monitoring, SEERTM 1000 made by GE Medical Systems Information Technologies, Inc, patients with ischemic stroke and normal 12-lead ECG tracing are assessed for paroxysmal atrial fibrillation (46).

As the Dutch organization for Cardiology recommends (47), before application of a Holter ECG monitor, the followings should be met, date of application, patient data, relevant medical use, and the existence of a pacemaker or an implantable cardioverter-defibrillator. Whenever a patient experiences symptoms, they should be timed and recorded. Changes throughout the day are also noticed by the patient, such as time of sleep, resting periods, and physical activity. Furthermore, the patient is informed about the basic guidelines for using Holter equipment (for example, avoid using an electric blanket and do not shower) and what to do if any of the electrodes become detached. A qualified cardiac physiologist analyzes and reports the Holter study; however, the supervising cardiologist is ultimately responsible for the Holter analyses’ correct authorization. Atrial fibrillation are defined as chaotic atrial arrhythmia with loss of p wave activity and irregular RR interval lasting for at least 30 seconds (48)

#### Echocardiography

Transthoracic Echocardiography (model Vivid ^TM^ T9 manufactured by GE Healthcare, USA 2018) are recommended only in selected patients with ischemic stroke and additional characteristics like evidence of cardiac disease on history, examination, or electrocardiogram (ECG), suspected cardiac source of embolism (example infarctions in multiple cerebral or systemic arterial territories), suspected aortic disease or paradoxical embolism and for patients with no other identifiable causes of stroke (49) Transthoracic Echocardiograms are read by cardiologists who are unaware of the patients’ neurologic condition or stroke subtype. Findings thought to be of potential diagnostic importance are extracted from the report view. Left atrial enlargement, patent foramen ovale (PFO)/atrial septal defect (ASD), low ejection fraction (EF35%), intracardiac thrombus, and valve vegetation or other valvular abnormalities being among them (50)

### Brain imaging

All patients undergo an acute CT scan SIEMENS (SOMATOM Definition Flash) to establish the stroke diagnosis and others undergo a brain MRI scan model MAGNETUM SPECTRA A TIM +Dot System 3T as part of a routine diagnostic workup. In addition, participants with stroke-like symptoms but negative CT scans for hemorrhagic stroke and uncertain ischemic stroke status are recruited for a study-specific MRI brain scan within the first 14 days after stroke. The MRI study protocol consists of 3D-T1, axial T2, 3D-FLAIR, DWI and SWI sequences. All patients will be evaluated for renal function status before undergoing brain imaging to reduce the risk of contrast-induced nephropathy (51)

The stroke volume (hematoma/infarct volume) is calculated using the ellipsoid method A+B+C/2, where A represents the largest diameter, B represents the largest diameter perpendicular to A, and C represents the product of slice thickness and number of slices. A, B, and C are measured in centimetres, and the resulting volume are expressed in cm3 or milliliters (8,52)

Leukoaraiosis are defined as bilateral areas of patchy or diffuse hypodensity on CT or white matter hyperintensity on MRI (Yang et al., 2015). The global brain atrophy is classified as not present if the width of the third ventricle is less than 5 mm, mild if the width is between 5 and 6 mm, moderate if the width is between 6 and 7 mm, and severe if the width is greater than 7 mm (53). All images are transferred to a computer workstation with an SYNGOVIA viewer and evaluated by two experienced radiologists.

#### Data management

Data are stored on password-protected and encrypted computers to maximize confidentiality and security. Each subject is assigned a unique, anonymous identifier linked to all collected data, including questionnaires and interviews. Data are identified only by ID numbers, and a link between names and numbers will be stored separately and safely. All paper files are kept in a secure, locked location accessible only to the research team. Data are only accessible to trained study personnel and locked away.

#### Safety consideration

At any point during the study, if a participant reports suicidal or self-harming thoughts, the research coordinator notifies the principal investigator immediately and the PI contacts the participant to conduct a risk assessment. As necessary, the PI initiates psychiatric consultations. Participants determined to require intervention receives information about available care resources, and those who do not need a higher level of care may continue to participate in the study and safety will always take precedence over participation in studies.

#### Data analysis

Data are entered on a Microsoft Excel sheet for statistical analysis and then converted to IBM SPSS PC version 26. Continuous variables are summarized as mean and standard deviation (SD), or Median and interquartile ranges, for non-parametric variables; frequencies and percentages will be used for categorical variables. Demographic variables, cardiovascular predictors of post-stroke cognitive impairment, stroke characteristics, and neuropsychiatric manifestations at one-month post-stroke will be compared between stroke survivors with cognitive impairment and without cognitive impairment using Chi-square in the case of categorical variables and Mann Whitney U test for non-parametric variables. Then, each post-stroke cognitive impairment predictor will be subjected to a univariate analysis using binary logistic regression; variables with p < 0.2 will be chosen for multivariate analysis (54). A multivariate Logistic analysis will be used to identify independent risk factors for post-stroke cognitive impairment at three months. At the same time, the odds ratio (OR) and the 95 % confidence interval (CI) will be determined. Statistical significance will be determined by a two-sided p ≤ 0.05.

#### Ethical issues

The Vice Chancellor’s office at the University of Dodoma provided permission to conduct the study after obtaining ethical clearance from the Directorate of Research and Publications (reference number MA.84/261/02). Following that, the administrative departments of Benjamin Mkapa and Dodoma Regional Referral Hospitals approved data collection, respectively. Participants are informed that their participation was fully optional and that they might opt out at any moment without affecting their daily routine care. To ensure privacy and confidentiality, participants’ names are substituted with identification numbers; however, their standard of care are unaffected by their decision to participate or not. Stroke survivors who had depressed symptoms are referred to a specialized psychiatric for additional e and treatment

#### Study timeline

The duration of this study is expected to takes 18-months to reach the required sample size. The first patient was recruited on the June 2020 and still on-going. Three months of follow-up is needed for each patient; however, overall data collection will need at least eighteen months..

## Discussion

Cross-sectional studies have investigated the relationship between cognitive impairment and other difficulties, such as disability and executive function limitations. Dependence, competency, and disability have been linked to cognitive impairment (55,56). Additionally, the cognitive decline between seven days and three months was associated with an approximately two fold increased risk of mortality compared to cognitive stability (57,58). The intervention for PSCI among stroke survivors which is debatable, as only cognitive rehabilitation improves cognitive impairment in stroke survivors, whereas there are no strong evidence for pharmacotherapy, lifestyle modifications, and cardiovascular risk factor interventions improve cognitive performance in stroke survivors (59)

This study has the potential to inform the burden and predictors of cognitive impairment among stroke survivors in our setting, considering the paucity of data in sub-Saharan Africa at large(60). However, as prospective cohort study is concerned, it is often difficult to maintain contact with all cohort members, mainly if the cohort is large and the length of follow-up is extensive. Loss of contact with many patients can lead to biased results, especially if the reason for loss to follow-up is related to the risk factor or outcome of interest. Furthermore, cohort study has limited capacity to infer causation from the observed association between stroke and cognitive impairment among survivors due to lingering confounding variables.

The final results will be submitted to the University of Dodoma library, the study sites (Dodoma regional referral hospital and the Benjamin Mkapa Hospital), and a manuscript will be ready for submission in different peer-review journals before publication.

## Data Availability

Data will be available and shared as per agreement of terms and conditions once the the data collection is completed.

## Authors’ contributions

Conceptualization: Azan Nyundo, Meda John

Data curation: Baraka Alphonce

Formal analysis: Ali Kinyaga

Funding acquisition: Tanzania Ministry of health

Investigation: Baraka Alphonce

Methodology: Baraka Alphonce, Azan Nyundo, Meda John

Project administration: Azan Nyundo, Meda John

Resources: Baraka Alphonce, Azan Nyundo, Meda John

Supervision: Azan Nyundo, Meda John

Writing – original draft: Baraka Alphonce,

Writing – review & editing: Azan Nyundo, Meda John

## Acknowledgments

First and far most, we acknowledge the patients and the staff of Dodoma regional hospital and Benjamin Mkapa hospital for their participation in the work.

## Appendix

### Aim 1

**Table 1:**
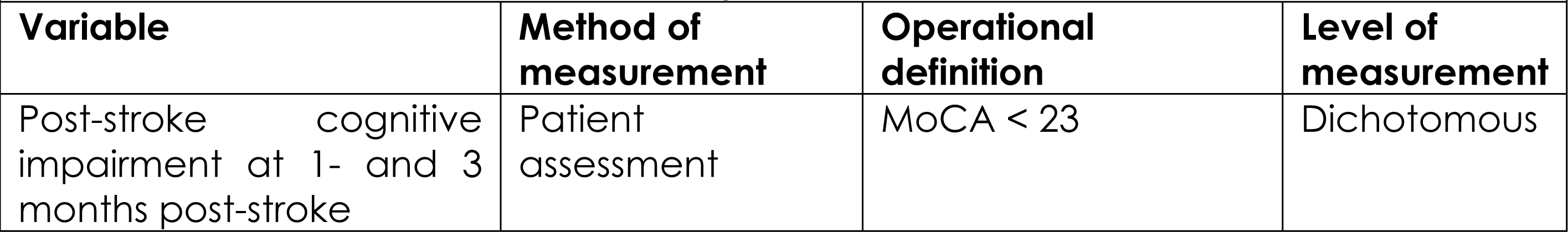
summary and description of objective number 1 variables of the study.

### Aim 2

**Table 2:**
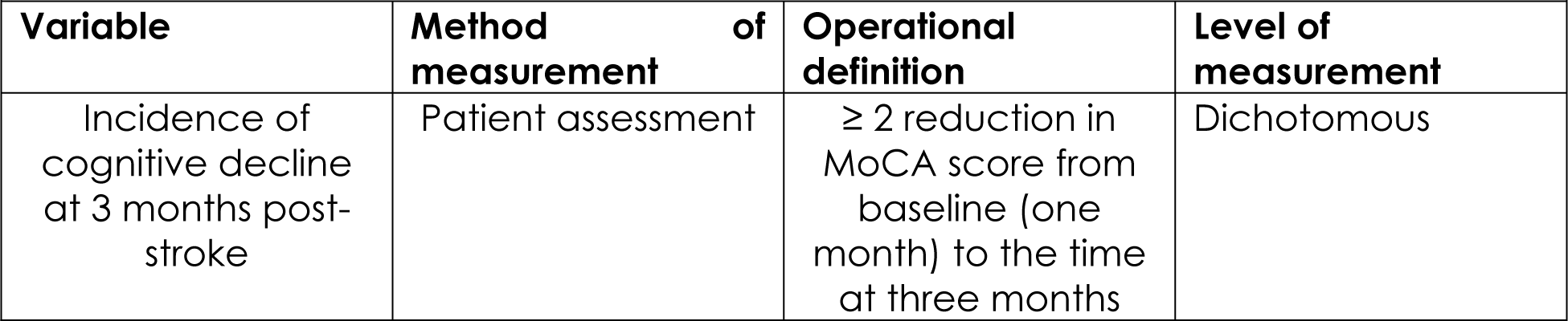
summary and description of objective number 1 variables of the study.

### Aim 3

**Table 3:**
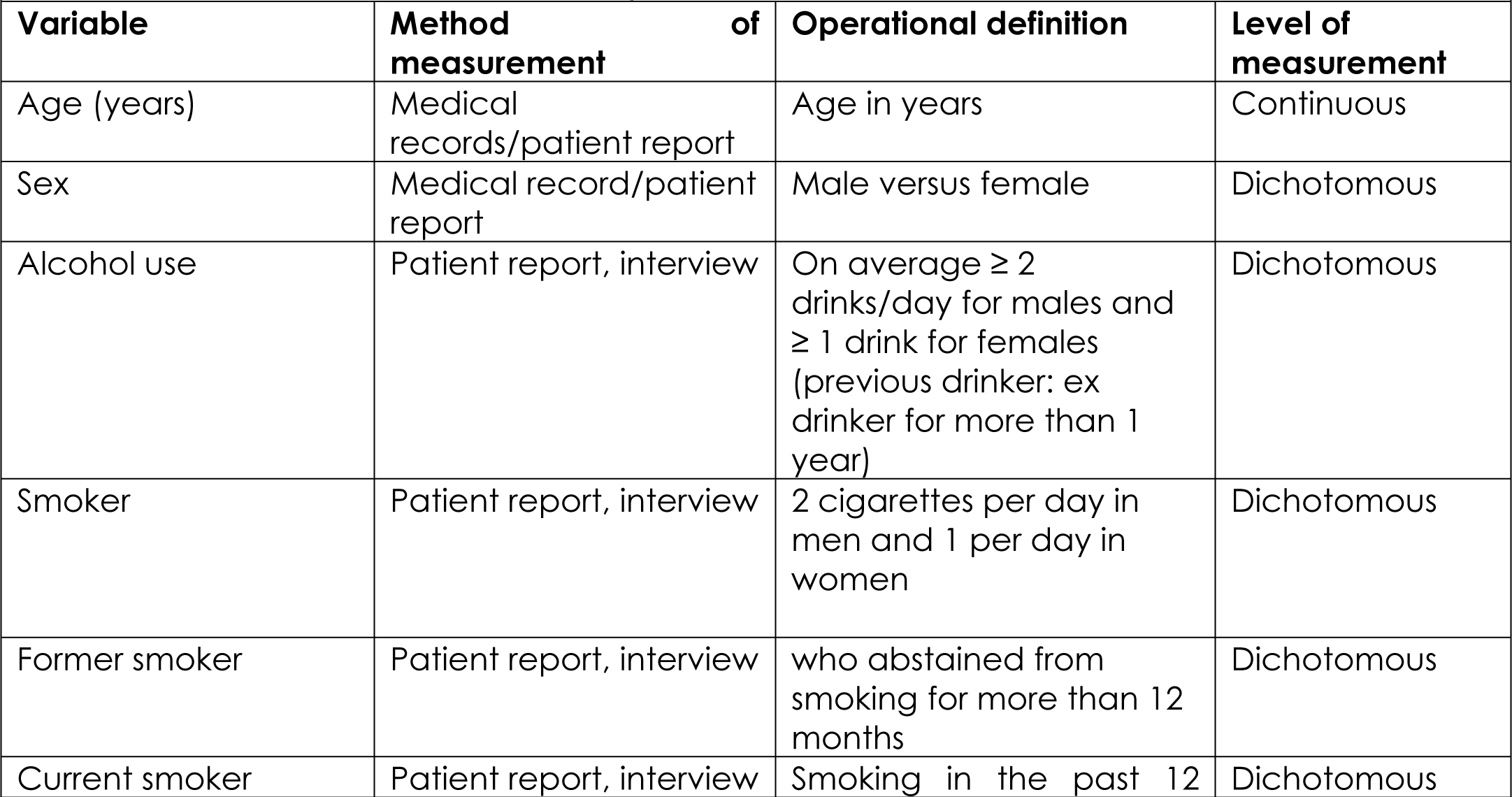
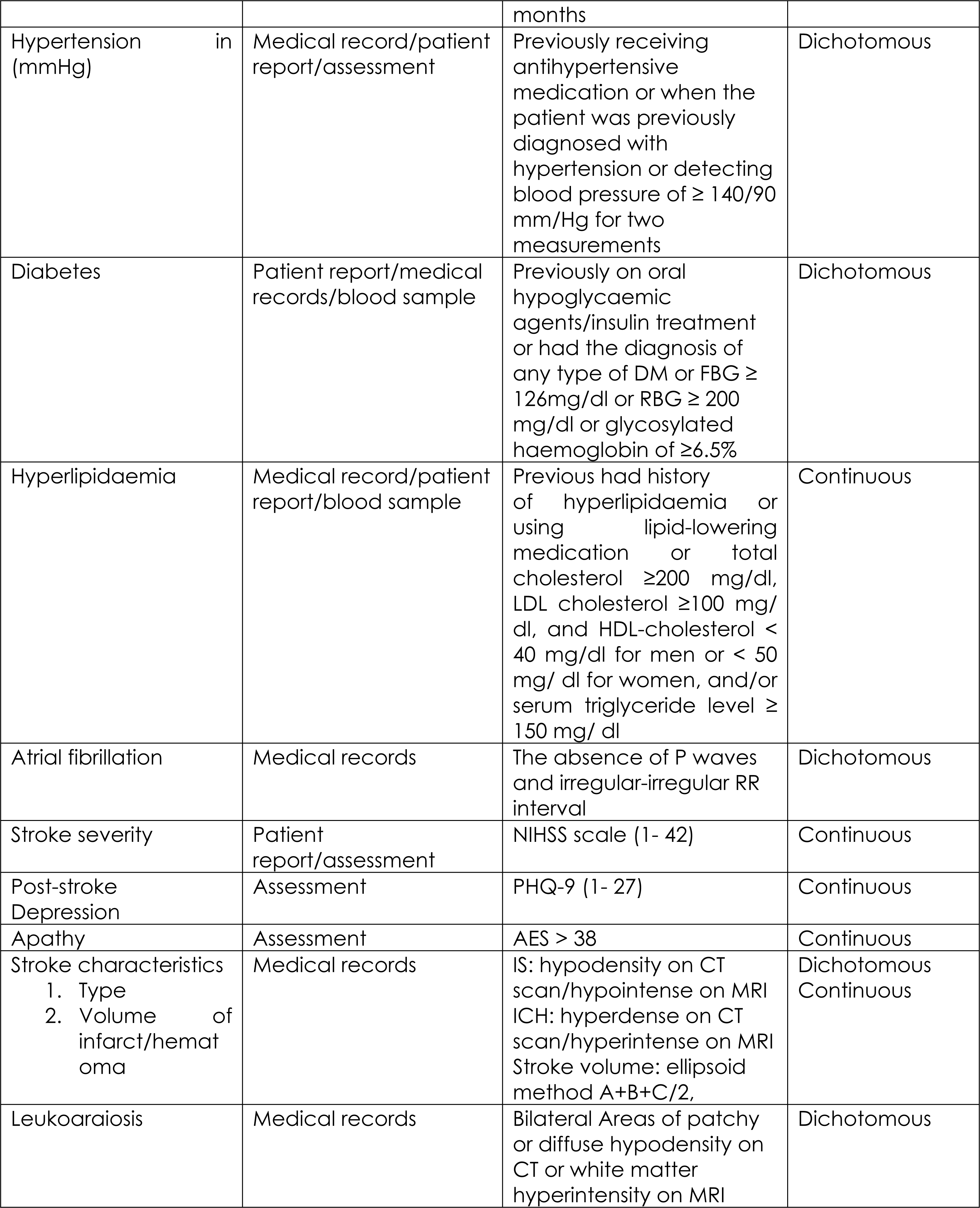
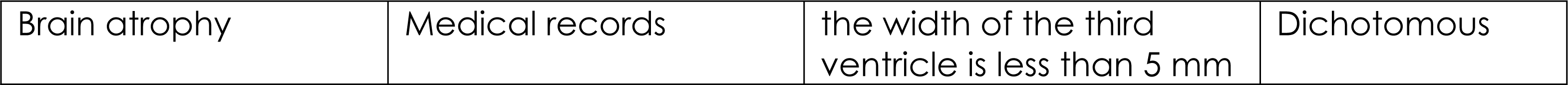
summary and description of objective number 3 variables of the study.

